# Explainable deep learning for disease activity prediction in chronic inflammatory joint diseases

**DOI:** 10.1101/2023.12.05.23299508

**Authors:** Cécile Trottet, Ahmed Allam, Aron N. Horvath, Axel Finckh, Thomas Hügle, Sabine Adler, Diego Kyburz, Raphael Micheroli, Michael Krauthammer, Caroline Ospelt

## Abstract

Analysing complex diseases such as chronic inflammatory joint diseases (CIJDs), where many factors influence the disease evolution over time, is a challenging task. CIJDs are rheumatic diseases that cause the immune system to attack healthy organs, mainly the joints. Different environmental, genetic and demographic factors affect disease development and progression. The Swiss Clinical Quality Management in Rheumatic Diseases (SCQM) Foundation maintains a national database of CIJDs documenting the disease management over time for 19’267 patients.

We propose the Disease Activity Score Network (DAS-Net), an explainable multi-task learning model trained on patients’ data with different arthritis subtypes, transforming longitudinal patient journeys into comparable representations and predicting multiple disease activity scores.

First, we built a modular model composed of feed-forward neural networks, long short-term memory networks and attention layers to process the heterogeneous patient histories and predict future disease activity.

Second, we investigated the utility of the model’s computed patient representations (latent embeddings) to identify patients with similar disease progression.

Third, we enhanced the explainability of our model by analysing the impact of different patient characteristics on disease progression and contrasted our model outcomes with medical expert knowledge. To this end, we explored multiple feature attribution methods including SHAP, attention attribution and feature weighting using case-based similarity.

Our model outperforms non-temporal neural network, tree-based, and naive static baselines in predicting future disease activity scores. To identify similar patients, a *k*-nearest neighbours regression algorithm applied to the model’s computed latent representations outperforms baseline strategies that use raw input features representation.

**Author summary:** Chronic inflammatory joint diseases affect about 200′000 patients in Switzerland alone. These conditions lead to immune system dysfunction resulting in inflammation that targets the joint tissues. Understanding which aspects of patients’ characteristics and disease management history are predictive of future disease activity is crucial to improving patients’ quality of life.

A significant obstacle to the widespread adoption of deep learning (DL) methods in healthcare is the challenge of understanding their “black-box” nature (i.e. the underlying decision process for outcome generation). Therefore, the development of “explainable” deep learning methods has become an active area of research. These approaches aim to provide insights into the inner workings of deep learning models, enabling physicians to understand and assess the output of DL models more effectively.

We propose DAS-Net: an explainable deep learning model that finds similar patients and predicts future disease activity based on past patient history. In our analysis, we contrast different explainability approaches highlighting which aspects of the patient history impact model predictions the most. Furthermore, we show how computed patient similarities allow us to rank different patient characteristics in terms of influence on disease progression and discuss how case-based explanations can enhance the transparency of deep learning solutions.

## 1 Introduction

Chronic inflammatory joint diseases (CIJDs) cause the immune system to attack healthy organs, particularly the joints [1]. In addition to causing pain, the inflammation can lead to synovitis, bone erosions, muscle and ligament damage. To this day, there exists no cure and the treatments primarily help attenuate the patients’ symptoms and improve their quality of life. Finding ways to minimise the disease activity is crucial to alleviate the disease burden on patients’ everyday life.

Digitalising patient healthcare data has led to a massive increase in available electronic health records (EHRs), opening up the opportunity to mine these records and employ machine learning (ML) approaches to discover novel evidence about real-world treatment efficacy and patient outcomes [2]. Due to the complex patient-specific disease progression patterns, CIJDs patient registries are very heterogeneous in the collected measurements and temporally sparse, presenting a challenge for ML models to learn from the data. In this work, we use the database of the Swiss Clinical Quality Management in Rheumatic Diseases (SCQM) Foundation [3]. It is a national longitudinal database of CIJDs documenting the disease management over time for 19’267 patients with different forms of arthritis.

We propose the Disease Activity Score Network (DAS-Net), an explainable multi-task neural network model to transform heterogeneous longitudinal patient journeys from the SCQM registry into comparable representations and predict future disease activity scores (DAS). DAS-Net evaluates the importance of the different aspects of individual management history (events) to predict future disease activity scores (i.e. multi-task forecasting). To this end, we trained our model on patients who had available DAS28-BSR (hereafter DAS28) [4] or ASDAS-CRP (hereafter ASDAS) [5] scores, without limiting our analysis to a specific arthritis subtype, but rather including all the patients for which either of these scores was available. The model is composed of multilayer perceptrons, long short-term memory networks [6], and augmented with attention mechanism [7] to process heterogeneous patient histories. The attention mechanism highlights parts of the patients’ histories that are most likely contributing to the outcome prediction, providing further insights into the model’s decision-making process.

Compared to physicians who use their experience to assess possible similarities among patients [8], we use our model to retrieve patients with similar disease progression by mapping the patients’ raw entangled data into a latent space with higher separability [9]. We empirically assessed DAS-Net’s ability to cluster patients with similar disease progressions.

Lastly, we explored multiple explainability approaches in our analysis, in particular through the (a) SHAP (SHapley Additive exPlanations) [10] value computation on the baseline models’ input features to gain post-hoc insights into the contribution of each feature (b) two-layered attention mechanism in the model architecture assigning weights to the different events of the patient histories and highlighting their significance for the model’s predictions, and (c) case-based importance weighting of the features for patient similarity assessment. We offer visual insights to illustrate how the model evaluates the similarity between some example patients and highlight the most influential features. To expand on these case-based explanations, we developed aggregate metric to rank the input features’ importance for similarity assessment.

By contrasting the results of these various approaches, we believe that we make a significant step towards enhancing the transparency of the model’s output.

### 1.1 Related work

Temporal deep learning models such as recurrent neural networks and transformers are commonly used in deep learning to analyse longitudinal patient data [11]. However, there is limited research on employing these temporal modeling approaches to predict disease progression in CIJDs. Most existing DL studies using CIJD databases focus on classifying the diagnoses rather than predicting how the disease progresses [2]. In studies that do predict disease progression, the continuous DAS values are usually simplified and thresholded into a binary classification task such as remission/no remission or response/no response, rather than predicted through regression [12]. For instance, Norgeot et al. [13] implemented RNNs to predict disease activity (remission/no remission) at the next rheumatology visit for rheumatoid arthritis patients. Their model significantly outperformed a static baseline, indicating the effectiveness of employing temporal models for modeling disease activity in CIJDs.

Furthermore, the majority of the existing studies are limited to patients with rheumatoid arthritis. However, in [14], both rheumatoid arthritis (RA) and axial spondyloarthritis (axSpa) patients were included and various non-temporal ML models (such as random forest, logistic regression and vanilla neural networks) were used to predict response/no response to different treatments. Their feature importance analysis revealed that different patient-reported outcome measures were the most significant predictors. This result supports our findings that past measures of disease activity are highly predictive of disease progression.

Our model architecture builds on the work proposed in [15] and further extends it (a) to support patients with different CIJD subtypes (not only RA) and (b) adding attention layers to measure the importance of different patient characteristics and management strategies for the model predictions. To the best of our knowledge, this is the only study emphasising patient similarity and explainability in modeling temporal disease progression in CIJDs.

## 2 Materials and methods

### 2.1 Dataset

#### 2.1.1 Description

The SCQM Foundation maintains a national database of inflammatory rheumatic diseases since 1997. The database documents the disease management over time for 19^*′*^267 patients through clinical measurements during the visits, demographics, prescribed medications and patient-reported outcome measures (database snapshot from 01.04.2022). Patients are diagnosed either with rheumatoid arthritis (RA), axial spondyloarthritis (axSpA), psoriatic arthritis (PsA) or undifferentiated arthritis (UA).

#### 2.1.2 Preprocessing

The SCQM database documents the management and disease evolution of the patients spanning several types of records and sources. We kept four distinct sources of information:

1. **Demographics (Dem)**: Non-temporal patient features such as date of birth or gender.
2. **Clinical measures (CM)**: Clinical measurements collected during a visit, such as DAS or weight.
3. **Medications (Med.)**: Features related to a prescribed medication and its duration (i.e. start or stop).
4. **Patient-reported outcome measure (PROM)**: Patient self-reported disease activity scores (such as RADAI score [16]).

While the demographics are static and only collected once, the clinical measures, medications and PROM are low-frequency time series. We refer to these as “time-related events”.

As preprocessing steps, we discarded patients with less than three CMs with distinct measurements of ASDAS or DAS28, or no medication information. We also discarded records with missing dates in the time-related data, and the clinical measures without either DAS28 or ASDAS. We selected the features used in [15], and additional ones based on availability and clinical relevance. We included the 90% most prescribed medications. After preprocessing, 10^*′*^589 patients (with a total of 79^*′*^872 clinical measures) and 31 features remained. The list of features is shown in appendix S1 Table. Dataframes and features and Figure 1 shows the distribution of the two DAS we used as predictive targets (i.e. outcomes). Summary statistics of the features are available in the tables of appendices 3.4, 3.5, 3.6, 3.7, 3.8, 3.9, 3.10, 3.11.

**Fig 1.**
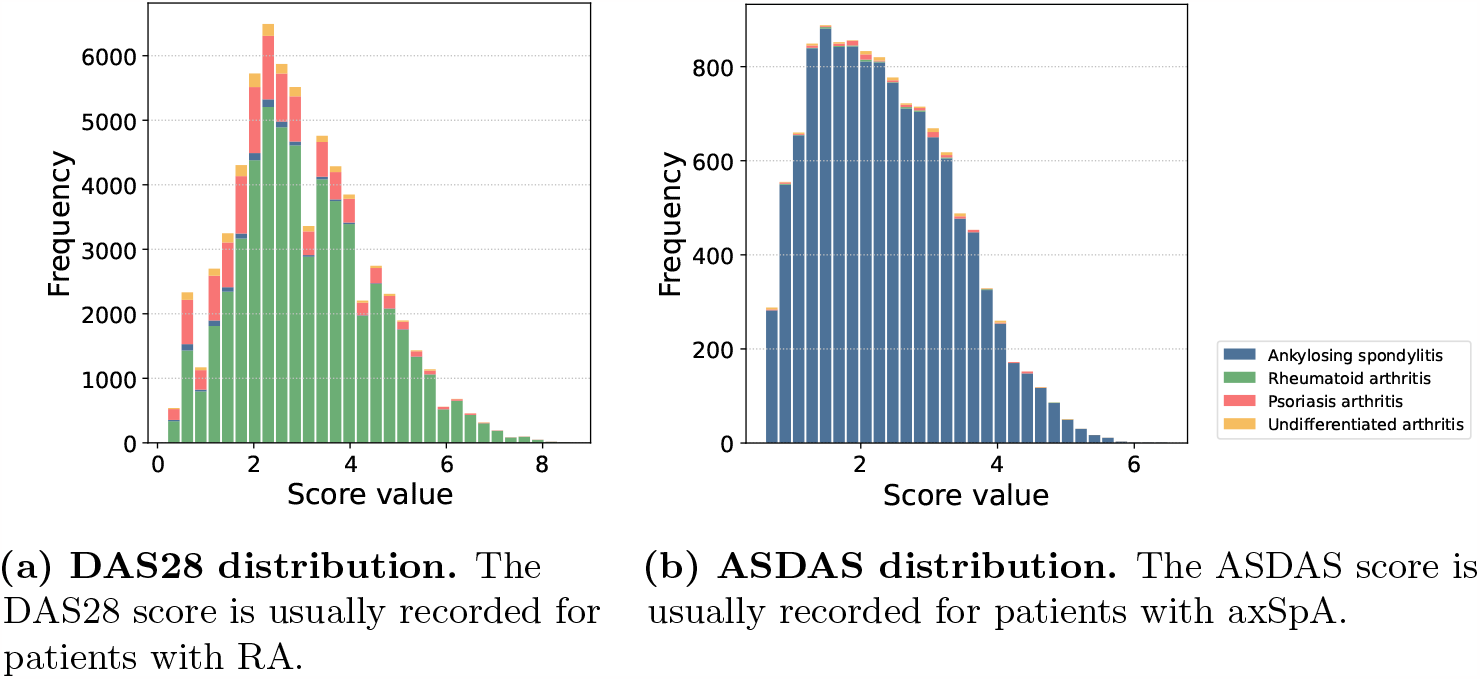
Disease activity scores distribution. Stacked histograms showing the DAS28 and ASDAS distribution in the preprocessed dataset. The different colour bars show the different arthritis types.

### 2.2 Model

#### 2.2.1 Motivation

Our dataset, like many EHR datasets, is irregular in both the temporal aspect (patients do not have the same number of medical visits), and in the number of recorded features (patients have varying numbers of recorded measurements and missing attributes).

Using non-temporal machine learning approaches (i.e. models that ignore patients’ full history) would limit the modeling of the data by restricting the input features to the subset shared by most data points or by discarding and imputing features to homogenise the data. This approach usually implies discarding most temporal information and using only the dataset’s main features, leading to significant information loss, poor generalisability and bias.

With this in mind, our goal is to develop a deep learning model that can process the full patients’ history, overcoming the challenges of temporal and feature irregularity. Moreover, it should be modular and support multiple outcome predictions allowing us to learn from all patients in the dataset with different DAS scores and arthritis subtypes. Lastly, it should produce meaningful latent representations, allowing us to compare patients with heterogeneous histories. An overview of the project pipeline, from data collection to implementation and evaluation of the different models is provided in Figure 2.

**Fig 2.**
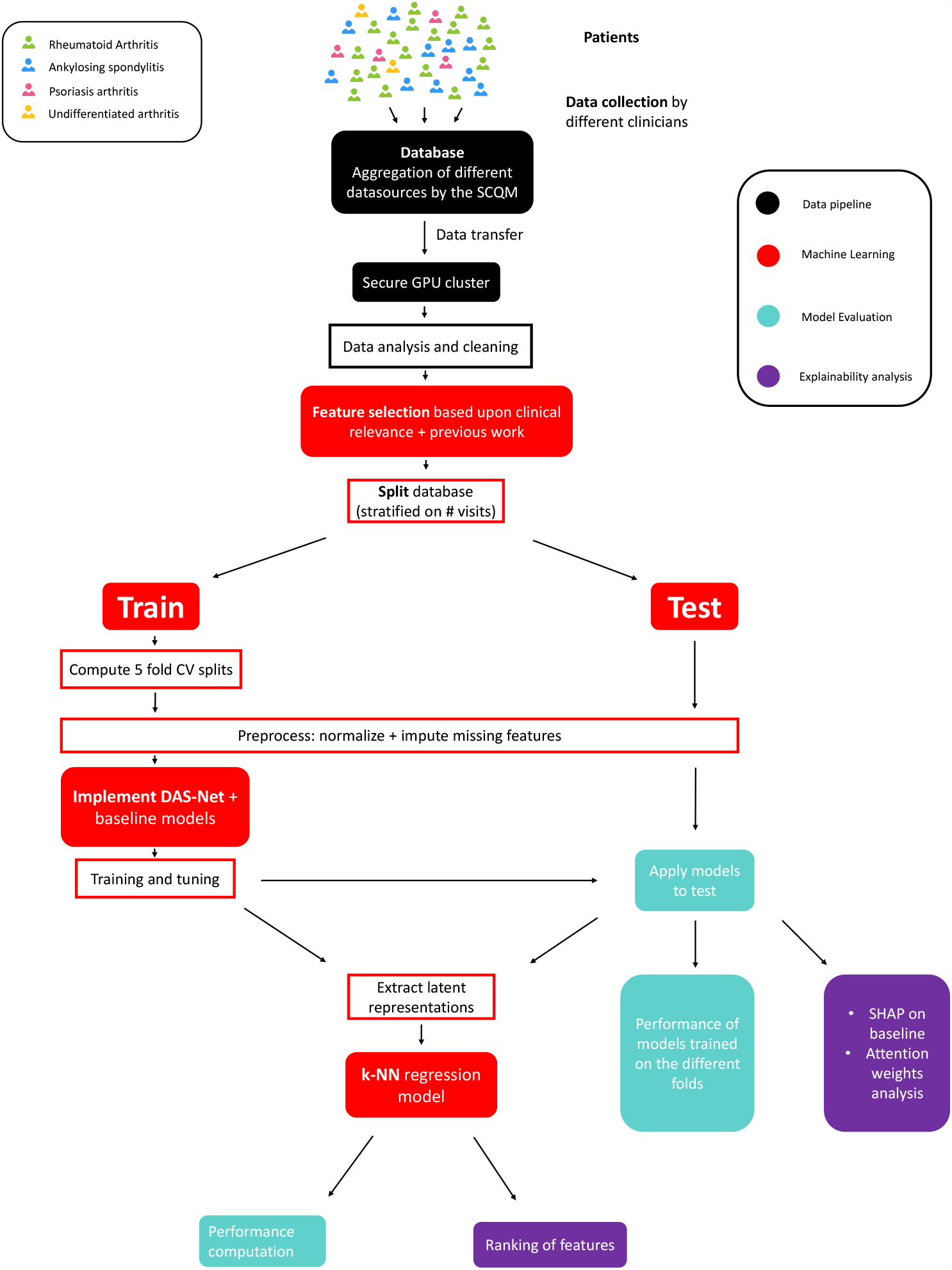
Project pipeline from data collection to implementation and evaluation of the different models.

#### 2.2.2 Architecture

Our model combines two main components. First, the model uses multilayer perceptrons (MLPs), long short-term memory networks (LSTMs) [6] and is augmented with attention layers [17] to build explainable vectorised patient representations. The different sources of information in the patient histories are handled separately until aggregation in the representation block. Then, we trained multilayer perceptrons to predict future DAS from these representations.

We adapted the architecture proposed in [15] to our setting by training multiple LSTMs, prediction networks, and by augmenting the model with several layers of attention layers. Figure 3 shows the model architecture with brief description for each component of the model.

**Fig 3.**
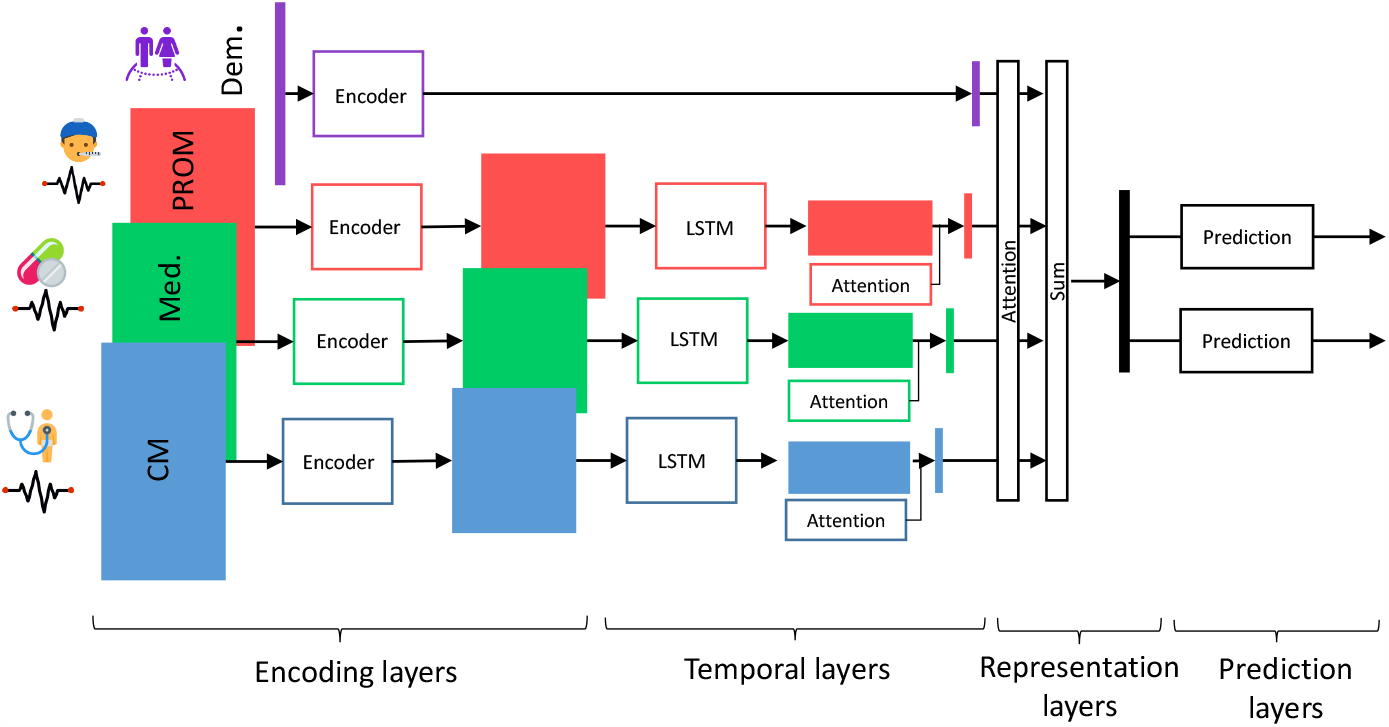
Model architecture. The encoders and prediction networks are MLPs. The model uses LSTMs to aggregate input sequences of different lengths and attention mechanism to weigh the different components of the input. “CM” stands for “Clinical measures”, “Med.” for “Medications”, “PROM” for “Patient-reported outcome measure” and “Dem.” for “Demographics”.

##### Model input

The input features are the patient medications, PROM and CMs up to a chosen time point, the demographics and the time to the prediction. Demographics, medications, PROM and CM are treated separately since their measurements are not aligned in time and contain different features. Merging them would result in a very sparse matrix and necessitate significant feature imputation.

##### Model output

The model predicts the next available DAS28 or ASDAS score by feeding the computed latent representation in the penultimate layers (i.e. representation layers) to two separate blocks of prediction layers. The latent representation is used posthoc to compute patient similarities.

##### Encoders

First, the MLP encoders process the normalised event-specific features. We defined separate encoders for each type of information (CM, Dem, PROM and Med). The encoders output lower dimensional embeddings for the time-related events and higher dimensional embeddings for the demographics to have matching history sizes in the later aggregation step. The order of the initial events is maintained in the computed embeddings.

We describe how the model is applied to a patient *p*. Let *ev* ∈ {*CM, Med, PROM*} be a time-related event, *s*_*ev*_ the number of features for *ev, q*_*ev*_ the embedding size, 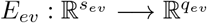 be the corresponding encoder and 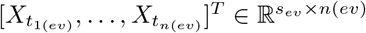 the ordered events measured at times 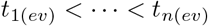. To ease the notation, we omitted the dependencies to *p*. We store the time-ordered embeddings 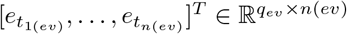 with 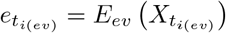.

For the demographics event, we simply have *e*_*dem*_ = *E*_*dem*_(*X*_*dem*_), where 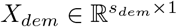 are the demographic features.

##### Temporal block

For a given sequence of events, the temporal block aggregates the embeddings into a one-dimensional vector. It contains one LSTM and one attention mechanism per category of time-related events. The LSTMs process the ordered embeddings computed by the event encoders. The attention mechanism is a trainable vector that weighs the contribution of each output of the LSTMs to the aggregated event history. For a given event, the aggregated history vector is the weighted sum of the outputs of the LSTM.

Thus, let *L*_*ev*_ be the LSTM for event *ev, ev* ∈ {*CM, Med, PROM* }. *L*_*ev*_ takes as input the sequence of embeddings 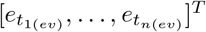 and outputs a processed sequence 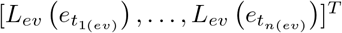. Given the computed local attention weights 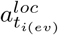, *i* = 1, …, *n*, the aggregated event history is

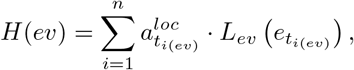

where using the *softmax* operator we have that 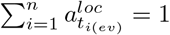.

##### Representation block

The representation block combines the event-specific outputs *H*(*ev*) of the temporal block, the demographics embedding *e*_*dem*_ and the time to prediction *t* into a unique vector. It is augmented by an attention mechanism, weighing the contribution of each type of event to the representation. The representation of a patient is the weighted sum of the demographics embedding and the aggregated event-specific histories, concatenated with the prediction time *t*.

Thus, *R* = [*P, t*] where

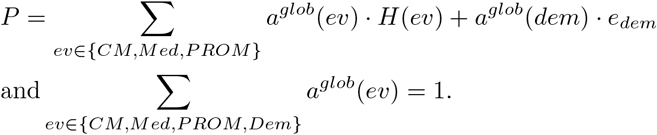

*R* = [*P, t*] is the combined latent representation of the patient history. It is used as input to predict future disease states and to compute similarities between patients.

##### Prediction networks

We defined two multilayer perceptron prediction networks, *P*_*DAS*28_ : ℝ^*r×*1^ → ℝ and *P*_*ASDAS*_ : ℝ^*r×*1^ → ℝ. The networks take as input the patient representation *R* and output the predicted DAS value at the medical visit at time *t*.

#### 2.2.3 Features and target selection

As described in subsubsection 2.1.2, we only included patients with at least three measurements of either DAS28 or ASDAS. These two DAS are part of the clinical measures, i.e. they are recorded during the medical visits of the patients. We use as targets the DAS collected from the second CM onwards, to ensure sufficient history length. The DAS from past CMs are part of the input features; a DAS is thus the target and then a feature once it becomes part of the patient’s history.

For each possible target, we used as input features the demographics and all the time-related events observed at least 15 days before the target CM.

#### 2.2.4 Optimisation

We stratified the patients on the number of CMs and randomly sampled 20% of the stratified patients as testing set that was not used for model training and tuning. We standardised the features and imputed missing values. We performed a five-fold CV on the training data to find the optimal parameters via random search. We selected the hyperparameters with the lowest average loss across the folds on their respective validation sets.

Following the empirical risk minimisation principle, our training objective is the sum of the mean squared error (MSE) for the DAS28 and ASDAS predictions. We used the AdamW [18] algorithm with mini-batch processing to optimise the objective.

At each step, we randomly sampled two batches of patients, one containing the patients with available DAS28 and the other with available ASDAS to ensure consistent joint optimisation of both objectives for these patients. We predicted all the available targets for each selected patient. The loss optimised at each optimiser step is defined in Equation 1

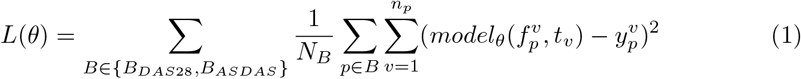

where *B*_*DAS*28_ and *B*_*ASDAS*_ are the sampled batches patients with available DAS28 and ASDAS respectively, *N*_*B*_ is the total number of targets in batch *B, n*_*p*_ is the number of targets for patient *p*, 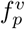 are the input features for patient *p* to predict target *v, t*_*v*_ is the time to target *v* and 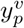is the true value of the target. *θ* denotes the model parameters to be optimised. We used batch sizes proportional to the total number of available targets per score to ensure consistent joint optimisation of both prediction networks.

### 2.3 Patient similarity: *k*−NN regression model

We evaluated the utility of DAS-Net’s computed latent representations (i.e. computed vector representation *R* as described in subsubsection 2.2.2) to retrieve similar patients. Given a patient representation at a prediction time-point, we computed the *L*^1^ distance to all other representations and selected the *k* closest patient embeddings (*k* = 50).

We matched the computed patient representations from the test set to their closest representations in the train set, such that for each patient representation *e*_*p,t*_ ≔ *e* ∈ *ℛ*_*test*_ (i.e. the computed representation embedding for patient *p* at time *t*), we found the subset of nearest neighbour representations *𝒩*_*e*_ ∈ *ℛ*_*train*_. We omitted the dependencies to *p* and *t* to ease the notation. This experiment simulates comparing incoming data to an extensive established database, possibly across hospitals. It could help find optimal management strategies faster by assessing which strategy worked best for similar patients.

Analogous to *k*−NN regression, we compared the representation’s future DAS with the average DAS of their closest matched set. We refer to this model as the *k*−NN regression model.

#### 2.3.1 Feature importance for similarity assessment

We developed aggregate metrics to assess the average importance given to each feature for the similarity computation between an index patient and their subset of nearest neighbours.

For continuous features, we computed the average absolute distance (AAD) between the feature value of the patients in the test set and the average value in their matched set (in the training data), and the standardised AAD by dividing the AAD by the standard deviation of the feature:

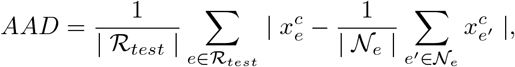

where 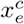 is the value of the continuous feature *c* for patient embedding *e*. For all computations, we restricted the subsets to the embeddings with available feature *c*. This metric reflects how much the values of the features of the subset of nearest neighbours deviate from the values of the index patient.

For a categorical feature *f*_*j*_ with possible categories *S*_*j*_ we computed the prior empirical probability of each category *k ∈ S*_*j*_. Furthermore, for each *k ∈ S*_*j*_, we computed the adjusted probabilities for the embeddings in the neighbourhood *𝒩*_*e*_ of an index patient embedding *e* with feature value *k*, i.e. the probability 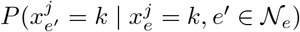. We compared the two quantities to evaluate the importance of each categorical feature for the similarity computation. For an embedding *e*^*′*^ *∈ ℛ*_*train*_, the prior empirical probability 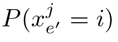of category *i ∈ S*_*j*_ is

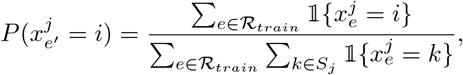

and the adjusted probability is

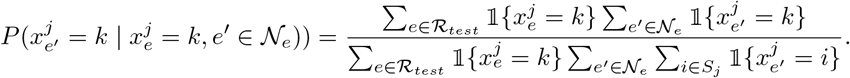

Again, we restricted the computations to the subsets of patients with available feature *j*. The increase in adjusted probabilities versus prior probabilities reflects how likely the feature is to have the same value as the index patient within its subset of nearest neighbours.

## 3 Results and Discussion

We compared the performance of DAS-Net and of the *k*−NN regression model for future disease activity prediction to different baseline models and further explored the three explainability approaches to better understand the relationship between input features and model output at different stages of the modeling process.

### 3.1 Performance

#### 3.1.1 DAS-Net prediction

We compared the performance of our model to two machine learning models: vanilla neural network (MLP), tree-based gradient boosting model (XGBoost), and one static naive baseline. The static naive baseline uses the last available DAS28 (resp. ASDAS) score for the given patient as its prediction. This strategy implies using the last disease state of a patient as a predictor of their future disease state. The MLP and XGBoost baselines take as input the same features as our model but only their last available values. Restricting the number of values per feature is necessary since these models cannot handle varying input sizes. We trained one MLP and XGBoost model per prediction task.

In Table 1 we report the models’ average performance and standard deviation on the test set. Our model achieves the lowest mean squared error (MSE) on both prediction tasks (MSEs of 0.510 *±* 0.009 for ASDAS and 0.965 *±* 0.014 for DAS28). In second place comes the XGBoost model performing the best out of all baseline models (MSEs of 0.534 *±* 0.003 for ASDAS and 0.992 *±* 0.002 for DAS28). Using a naive model that uses the most recent DAS score as prediction achieves the worst performance (MSEs of 0.842 for ASDAS and 1.475 for DAS28).

**Table 1.**
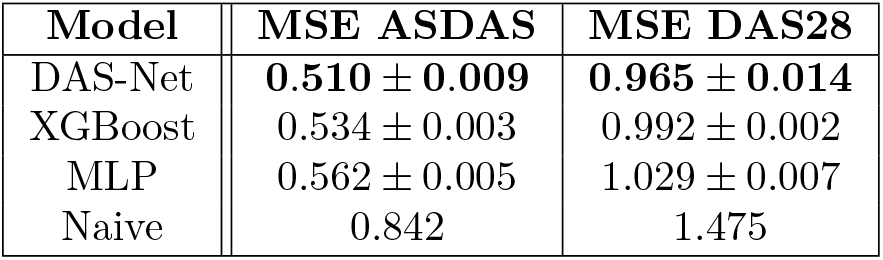
Model performance (regression). DAS-Net outperforms the three baselines for both prediction tasks. The naive baseline simply reuses the last available DAS. The MLP and XGBoost baselines use the last available values of each feature as input and our model the whole patient history.

Furthermore, we evaluated the models’ ability to correctly predict active RA (i.e. DAS28 values above 2.6) and moderate axSpA (i.e. ASDAS values above 2.0). To perform the classification, we trained a logistic regression model on DAS Net’s latent embeddings from the training set and evaluated the performance on the test set. We compared the performance of this approach to the XGBoost and MLP predictions, where we thresholded the predicted values of DAS28/ASDAS. Our approach achieves overall a higher accuracy than the baseline ML models (accuracies of 0.761 *±* 0.001 for ASDAS and 0.757 *±* 0.000 for DAS28 for our approach) (Table 2). Furthermore, the sensitivity and specificity of our approach are more balanced than for the baseline models. The baseline models achieve a higher sensitivity but suffer from a low specificity (Table 2).

**Table 2.**
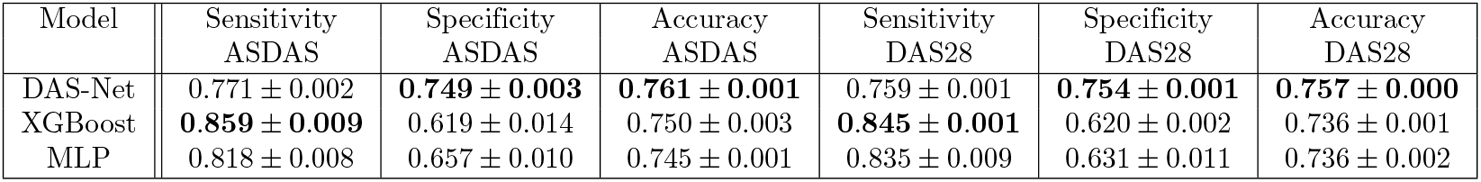
Model performance (classification). We evaluated the performance of the different approaches at predicting active disease (i.e. DAS28 values above 2.6 or ASDAS values above 2.0). While our approach has a slightly lower sensitivity than the baselines, it has a better balance between sensitivity and specificity and has an overall higher accuracy.

| Model | Sensitivity<br>ASDAS | Specificity<br>ASDAS | Accuracy<br>ASDAS | Sensitivity<br>DAS28 | Specificity<br>DAS28 | Accuracy<br>DAS28 |
| --- | --- | --- | --- | --- | --- | --- |
| DAS-Net | 0.771 ± 0.002 | **0.749 ± 0.003** | **0.761 ± 0.001** | 0.759 ± 0.001 | **0.754 ± 0.001** | **0.757 ± 0.000** |
| XGBoost | **0.859 ± 0.009** | 0.619 ± 0.014 | 0.750 ± 0.003 | **0.845 ± 0.001** | 0.620 ± 0.002 | 0.736 ± 0.001 |
| MLP | 0.818 ± 0.008 | 0.657 ± 0.010 | 0.745 ± 0.001 | 0.835 ± 0.009 | 0.631 ± 0.011 | 0.736 ± 0.002 |

**Table 3.** Similarity matching. The *k*-NN (*k* = 50) method based on the model latent embeddings outperforms the *k*−NN algorithm directly applied to the raw data and the completely random subset for the retrieval of similar patients.

| Model | MSE ASDAS | MSE DAS28 |
| --- | --- | --- |
| $k$-NN model on DAS-Net latent representations | **0.506** | **0.966** |
| $k$-NN on raw data | 0.681 | 1.218 |
| Random subset | 0.915 | 1.863 |

To understand the effect of the length of patient history on the prediction performance, we computed the model’s performance as a function of varying lengths of patient histories. Figure 4 shows the MSE decreases as more prior medical visits become available to the model. Additionally, in Figure 5, we plot the predicted versus ground truth DAS28 and ASDAS scores for two example patients, showcasing how DAS Net could be used by clinicians to monitor and predict disease activity.

**Fig 4.**
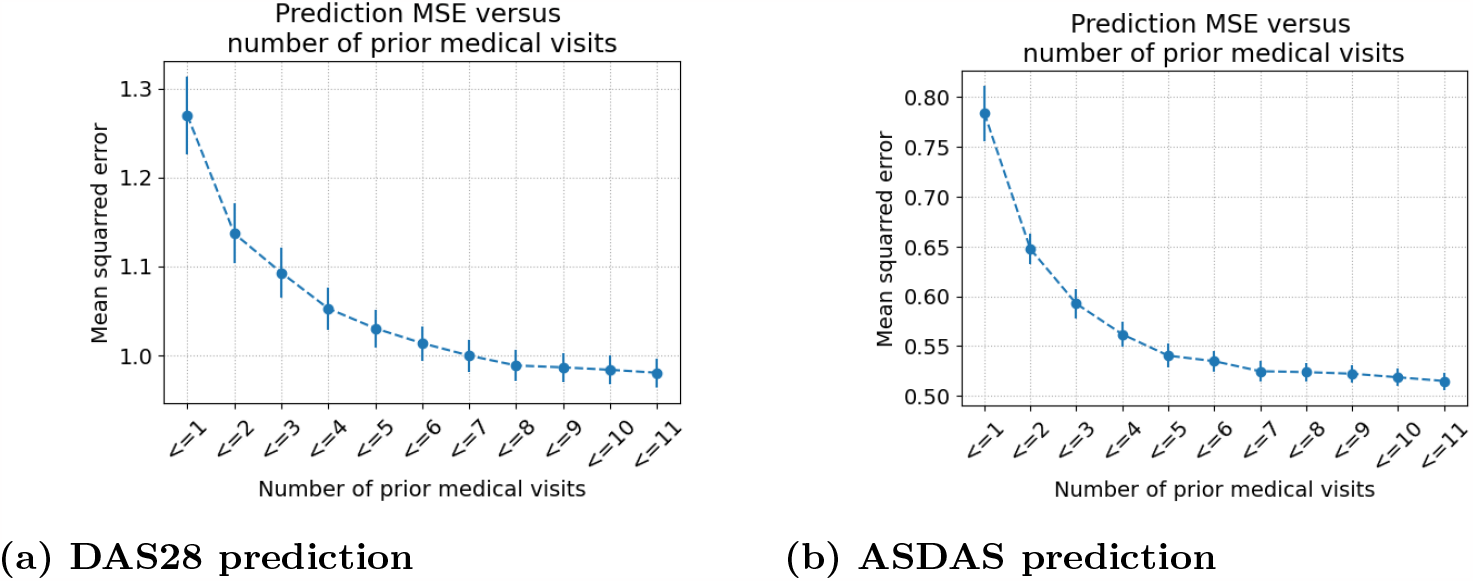
MSE versus number of prior medical visits. The MSE between model predictions and target DAS values decreases as the number of prior medical visits increases. The availability of at least three prior medical visits induces a steep decrease in MSE.

**Fig 5.**
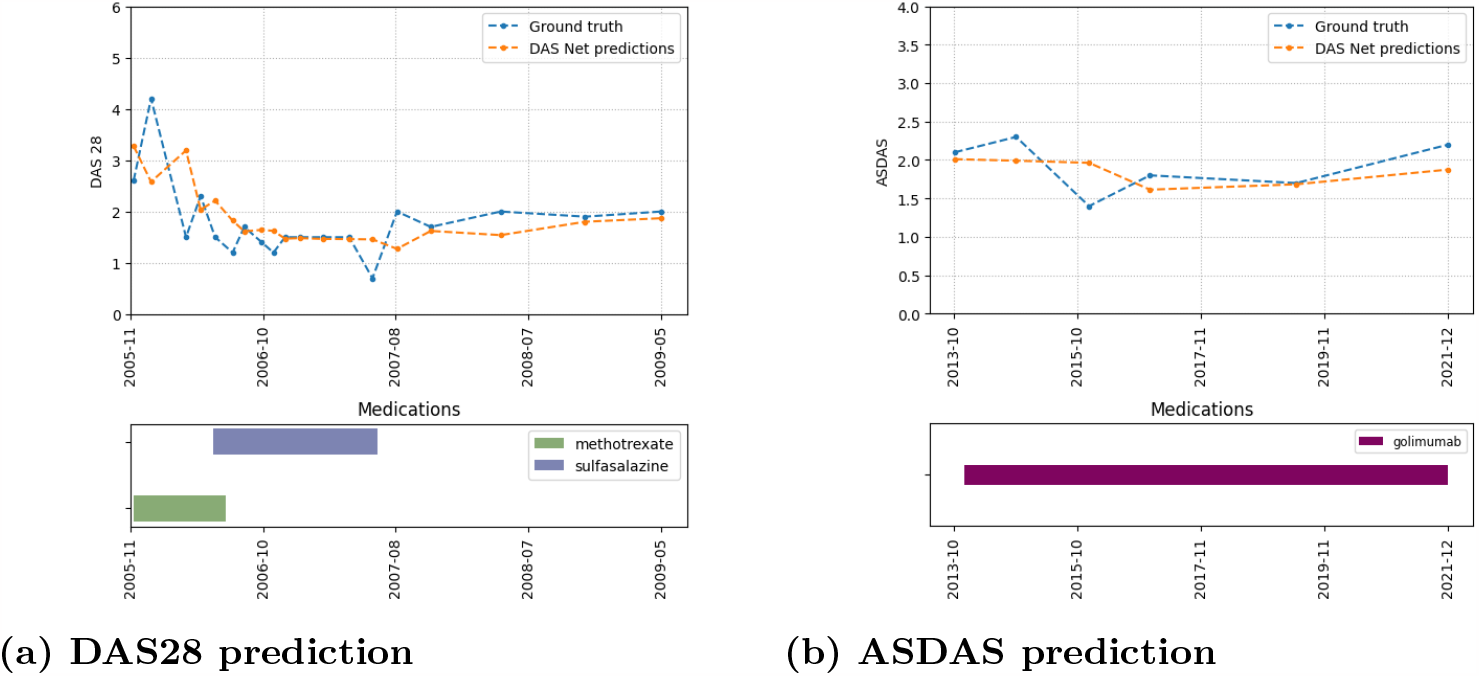
Predictions of individual patient trajectories. We compare the model predictions with the ground truth values of DAS28/ASDAS for two example patients. The bar charts show the prescribed medications present in the database.

#### 3.1.2 Patient similarity: *k*−NN regression model

We evaluated the ability of our model to cluster patients with similar disease progressions, by comparing the future DAS values of the embeddings in the test set with the average values of their most similar embeddings, as computed by our *k*−NN regression approach on DAS-Net’s latent embeddings. We compared the performance of our approach to the performance of a *k*−NN algorithm applied to the raw data, and a naive approach selecting a random subset of patients. Both baseline strategies thus do not utilise DAS-Net’s computed latent representations. The *k*-NN model on the latent representations achieves the lowest MSE (MSEs of 0.506 and 0.966 for ASDSAS and DAS28 prediction).

Interestingly, our *k*−NN approach has a similar predictive performance to the DAS-Net model for prediction (Table 1), and also outperforms the MLP and XGBoost baselines, suggesting that the DAS-Net latent representations successfully capture the important predictive components from the patient history.

### 3.2 Explainability approaches

In this section, we compare and contrast the results obtained from the different feature attribution techniques we applied or developed. These methods offer multiple insights on the relationship between input features and model output at different stages of the modeling processes.

#### 3.2.1 SHAP values on vanilla neural network

For the baseline neural network model (MLP), we computed the SHAP [10] values for the input features. SHAP values are derived from the game-theoretic-based Shapley values [19] and compute the contribution of each feature to the model predictions.

The plots in Figure 6 show the top-10 SHAP values for ASDAS and DAS28 predictions. Each dot represents a feature value from the test set and is overlaid with a colour reflecting the value of the feature. The x-axis shows the SHAP value. In our setting, a positive SHAP value indicates that the feature drives the model predictions upwards, and thus leads to higher predicted DAS. The features are ordered by the average magnitude of their SHAP values (from top to bottom, and we included only the top ten features). Overall, the SHAP values are consistent with the clinical knowledge.

**Fig 6.**
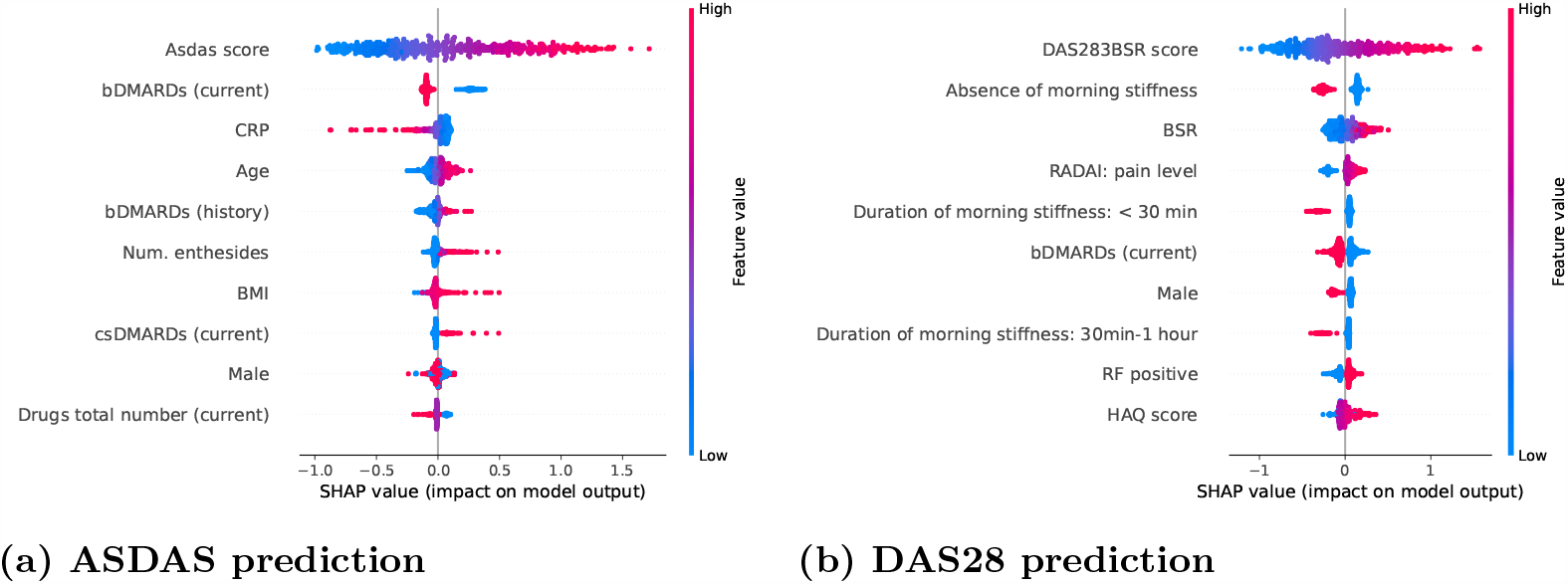
SHAP feature importance. The x-axis shows the SHAP value, and each dot is overlaid with a color representing the feature value. Thus, a pink dot with a positive SHAP indicates that the feature has a high value and leads to a higher predicted DAS. We show the top-10 features with the highest absolute SHAP values (ordered from top to bottom).

For ASDAS prediction, the *past ASDAS* values, *age* and *number of enthesitides* are positively correlated with their SHAP values, indicating that a higher value leads to a higher predicted disease activity score. For the medications, currently taking a *bDMARD* leads to lower future predicted DAS and the opposite for *csDMARDs*. For DAS28 prediction, the *past DAS28* values, *BSR, HAQ* and *RADAI pain level* are positively correlated with higher predicted disease activity scores. The *absence or short duration of morning stiffness* leads to lower predicted DAS. Being *male* is also a better prognostic factor.

Furthermore, we computed the absolute SHAP values of the features for each model trained on one of the 5 folds in our data (during 5-fold cross-validation). The plots in Figure 7 show the average and standard deviation of the absolute SHAP values for the 10 features with the largest overall absolute SHAP values (ordered from top to bottom). The importance ranking of the features is consistent across the different models.

**Fig 7.**
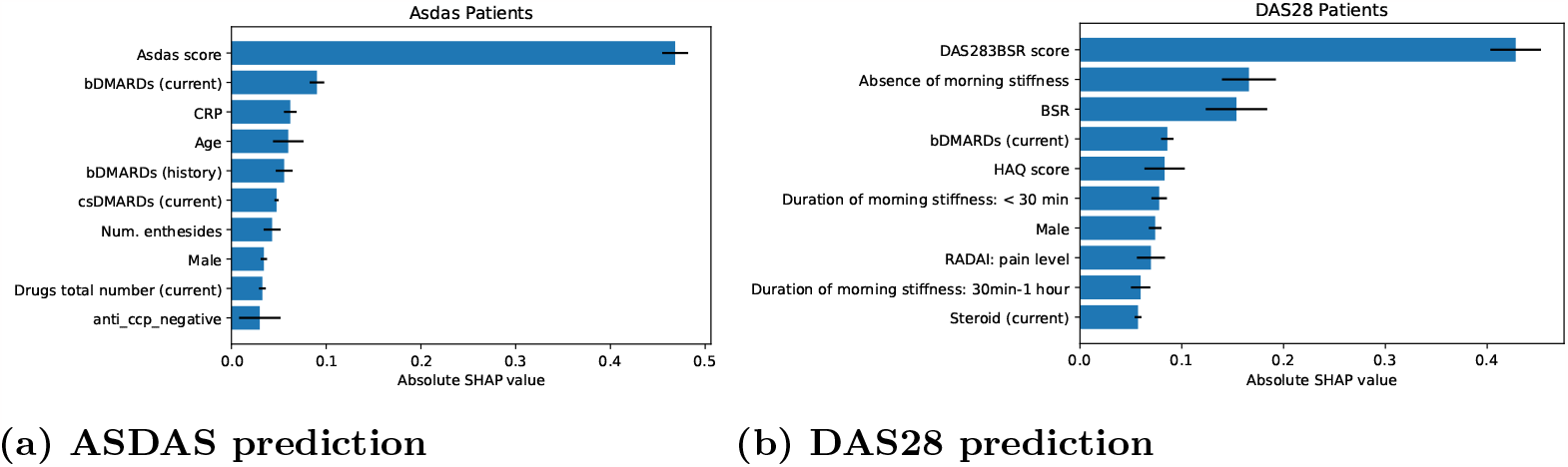
Mean and standard deviation of the absolute SHAP values across folds. We trained one MLP model per DAS on each cross-validation fold and computed their average absolute SHAP values on the test set. The top-10 most important features are consistent across folds.

##### Clinical relevance of findings

In predicting future DAS in RA patients, the model was strongly influenced by the presence and duration of morning stiffness, with no or shorter morning stiffness resulting in lower predicted DAS. Morning stiffness for more than one hour strongly correlates with DAS28 scores [20]. Thus, in the model, the level of morning stiffness might have reinforced the strong dependency of the future DAS from current and past DAS measurements.

Notably, the feature importance in predicting ASDAS in patients with axSpA differed with respect to the influence of current and past treatment. In RA, current use of bDMARDs predicted low DAS levels. Similarly, in axSpA, the current use of bDMARDs was linked to predicting low future disease activity. This suggests that bDMARDs are effective in managing disease progression in this context. However, in the axSpA cohort, the situation is more complex. Both past use of bDMARDs and current use of csDMARDs (conventional synthetic disease-modifying antirheumatic drugs) are connected to high future disease activity. This suggests that patients who have experienced previous failure with bDMARDs or require additional csDMARD therapy belong to a difficult-to-treat group with a low likelihood of responding favourably to future treatments.

#### 3.2.2 Attention weights

DAS-Net employs a two-layered attention mechanism for model-based explainability. The attention mechanism assign weights to the different events of the patient histories highlighting their significance for the model’s predictions. The **local attention** is specific to each type of time-related event showing the weight given to each event when building the aggregated event history (*H*(*ev*), *ev* ∈ {*CM, Med, PROM* } in subsubsection 2.2.2). For example, they show which specific clinical measure contributed the most to the prediction. The **global attention** gives weight to the aggregated event histories and demographics when building the patient’s full history representation (*P* in subsubsection 2.2.2). It shows which type of event is used the most by the model to make the prediction.

##### Global attention

Figure 8a shows the attribution of the global attention weights to the different event features (i.e. CM, PROM, etc.) in the patients’ history as the history length increases (denoted by the number of predicted targets). At the first target prediction, while most of the attention weight is already attributed to past CM, one-third is still attributed to other sources of information. Thus, when limited information is available, the model considers all the sources of information (i.e. clinical measures, medications, demographics and PROM). As the volume of available information increases (i.e. increasing length of history), the model increasingly assigns higher weights to the past clinical measures (CM) compared to the other sources of information. This weight distribution is reasonable because the previous CM contain the previous DAS that is predictive of future DAS.

**Fig 8.**
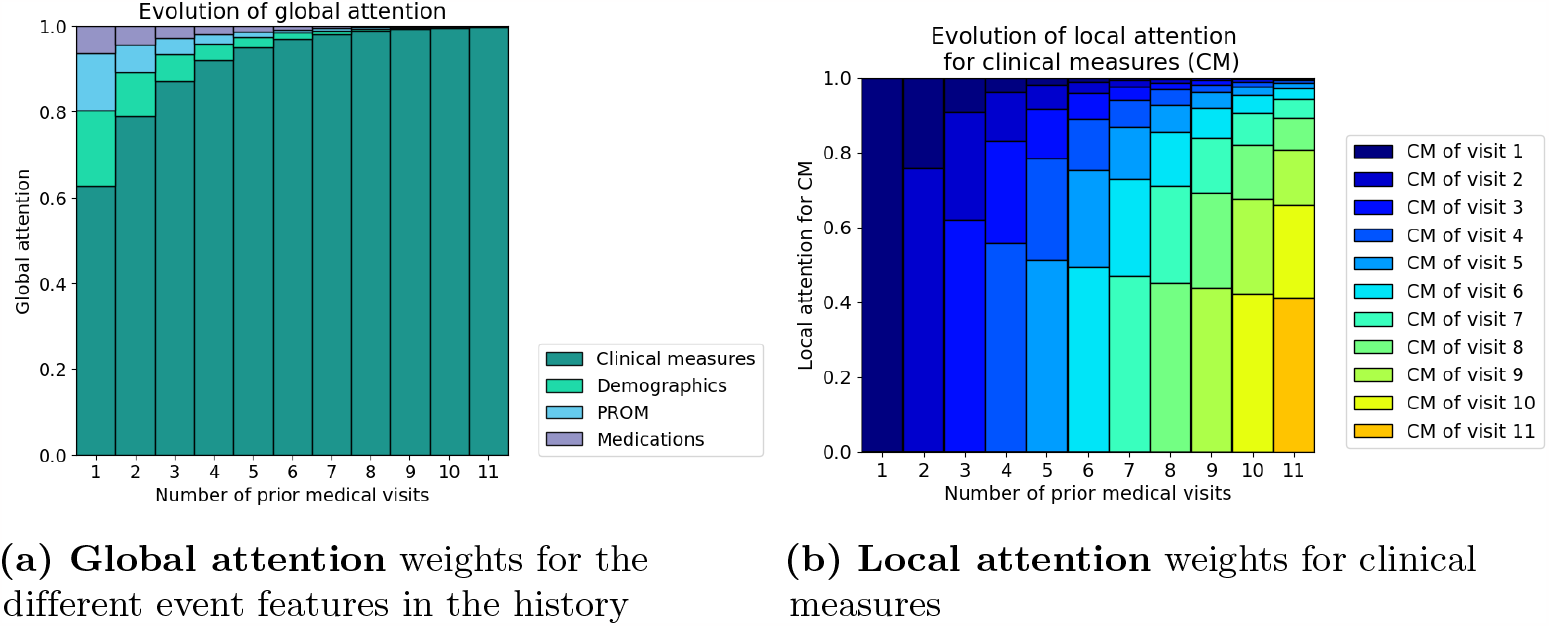
Local and global attention weights for increasing number of medical visits (i.e. increasing patient histories) aggregated over the patients in the test set. The **global attention** shows that the model uses clinical measures the most for the predictions. Furthermore, this pattern grows stronger as the number of available clinical measures increases. The **local attention** shows that within the clinical measures, most of the weight is attributed to the recent clinical measures.

Interestingly, for patients with a significant improvement in DAS (at least 20% improvement since the last CM), DAS-Net attributes less attention to the CM and redistributes it towards the other types of events (Figure 9).

**Fig 9.**
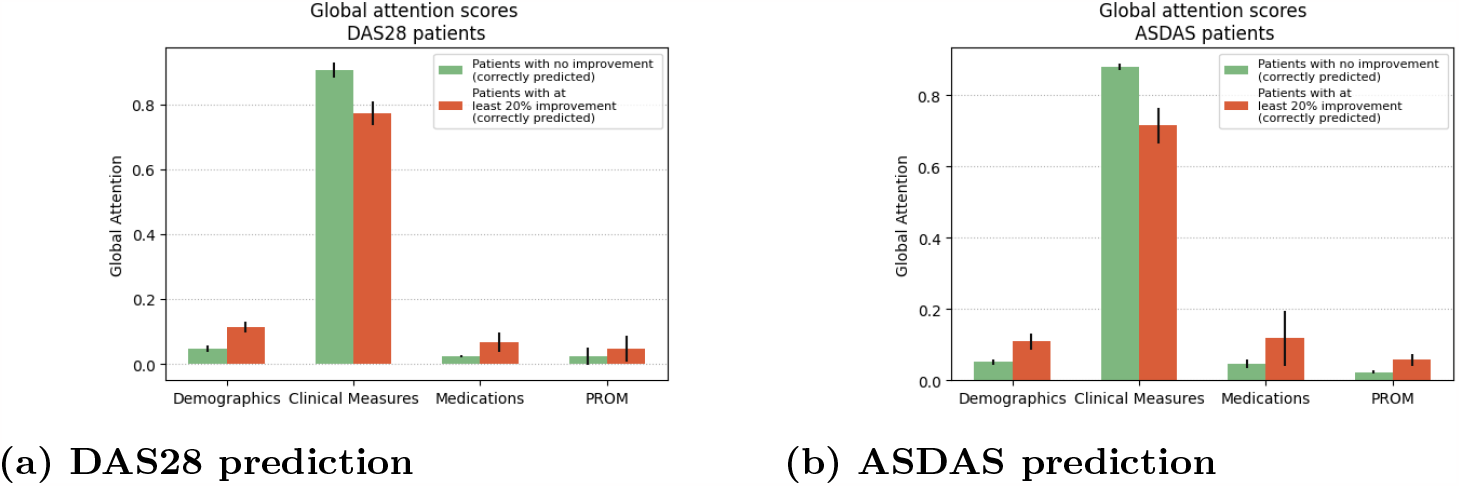
Global attention weights. Comparison in global attention weight attribution between patients with or without improvement in disease activity. The attention redistributes for patients with at least 20% improvement at the next visit.

##### Local attention

We further inspected the attribution of the local attention weights for the clinical measures in patients’ history when predicting the target outcome Figure 8b. Most attention is directed at the last available clinical measure in the history before the prediction. Furthermore, the attribution to past clinical measures is inversely proportional to their distance from the target. Our model thus assigns the highest attention scores to the recent clinical measures (i.e. latest measures), particularly the ones preceding the prediction.

#### 3.2.3 Patient similarity

##### Case-based visualisations

We visualised the patient representations by computing and plotting their two-dimensional t-SNE embeddings [21]. We plotted the embeddings for the entire cohort, i.e. the t-SNE embeddings of all the higher dimensional representations in *ℛ* = *ℛ*_*test*_ ∪ *ℛ*_*train*_. In Figure 10, we overlaid the embeddings in each subplot with colourmaps reflecting the values of the features. We reported the last available value for the given feature at the time of computation of an embedding (we restricted the plots to the embeddings with an available value for the feature). The subspace is separated according to different values of the features. In Figure 10a, we overlaid the embeddings with the CIJD subtype of the patients, even though this attribute is not explicitly used as an input feature in our model, to get an overview of the distribution of the different CIJD subtypes in the latent space.

**Fig 10.**
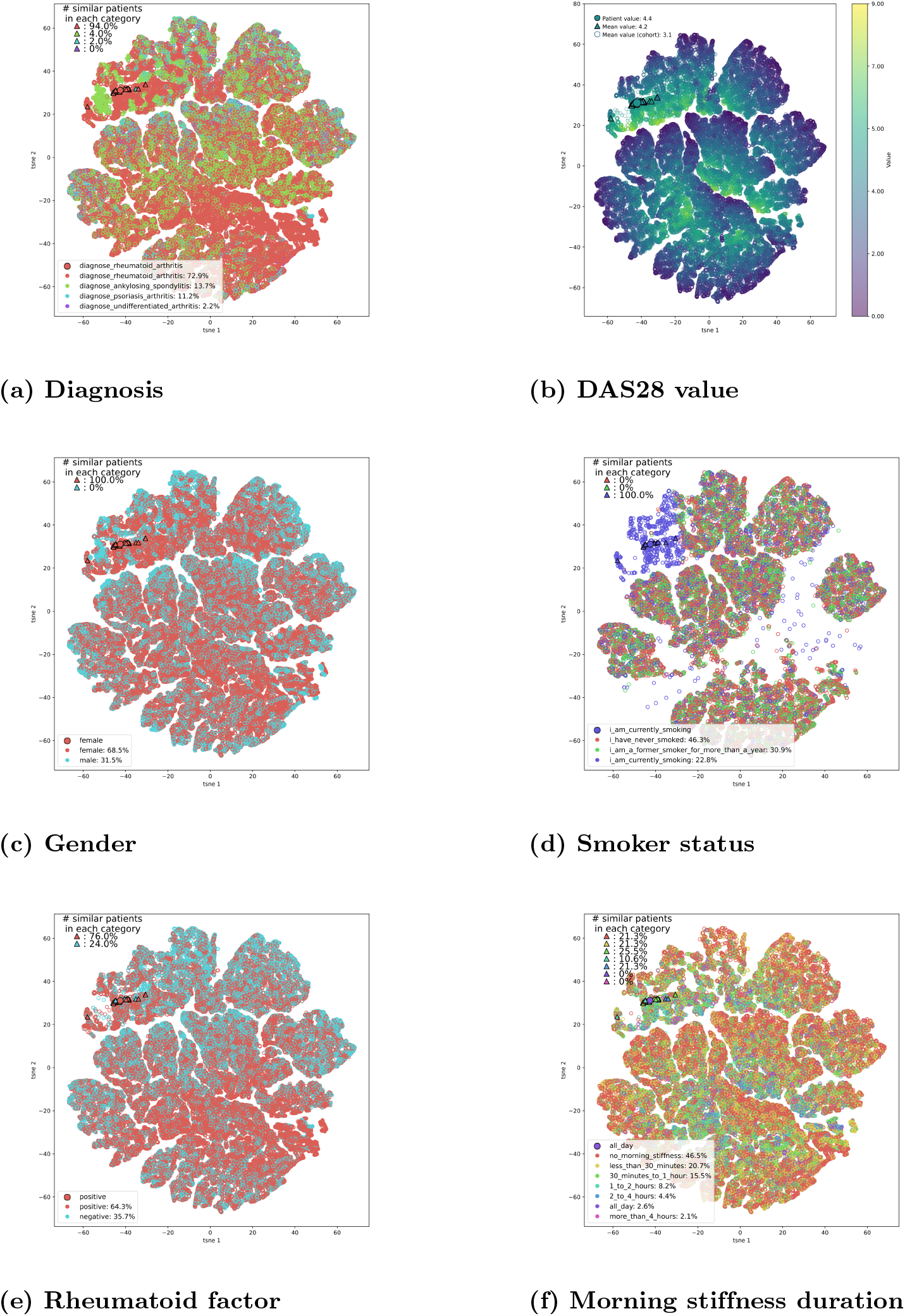
t-SNE visualisation of patient representations. Each point shows the t-SNE embedding of a representation of a patient at a given time. The subplots show the decomposition overlaid with the feature values (restricted to the embeddings with an available value for the feature). Furthermore, we highlighted a patient from the test set (larger filled dot) and her nearest neighbours (triangles) as computed by our algorithm. For each continuous feature we compute the average value in the entire cohort and within the subset of nearest neighbours. For categorical features, we computed the proportion of each category.

The plots provide general visual insight into the latent representation space. For instance Figure 10d shows the repartition of the smoker statuses, and a cluster of smoker patients in the top left of the figure stands out. Embeddings in this subspace correspond to patients with a smoking status that seems determinant for their disease activity prediction. Non-smoking patients and former smokers for more than a year are generally mapped to the same subspace, showing that the algorithm treats them the same. Some smokers, with possibly other more determinant factors, are also mapped in the same subspaces as non-smokers. By inspecting the gender plot (Figure 10c) we notice that males are generally mapped towards the edges of the sub-clusters. The same regions generally correspond to lower DAS28 activity regions (Figure 10b).

Furthermore, in Figure 10 we highlighted a randomly selected patient embedding *e*_*p,t*_ from the test set (larger dot) and its nearest neighbours (triangles) *𝒩*_*e*_ as computed by our *k*−NN regression model. For each continuous feature (here the *DAS28 score*) we also computed the average value in the entire representation set *ℛ* and within *𝒩*_*e*_. For categorical features (here *gender, duration of morning stiffness, rheumatoid factor* and *smoker status*), we computed the incidence of each category in *ℛ* and *𝒩*_*e*_. By comparing the overall distribution of the feature value with its distribution within *𝒩*_*e*_, we get insight into the importance given to the different features for the similarity assessment.

The example patient in Figure 10 is diagnosed with rheumatoid arthritis, and most of her nearest neighbours also belong to the same CIJD subtype (Figure 10a). She has a higher *DAS28 value* than average (4.4 versus mean cohort value of 3.1) and there is a distribution shift within her subset of nearest neighbours towards higher DAS28 values (average of 4.2 within her subset of nearest neighbours) (Figure 10b). Her *smoker status* (Figure 10d) and *gender* (Figure 10c) seem determinant for the similarity assessment, since all of her nearest neighbours are also smoking females. Conversely, the *rheumatoid factor* (positive Figure 10e) and *duration of the morning stiffness* (all day, Figure 10f) seem to be considered less important for this patient. However, there is still an overall redistribution towards positive rheumatoid factor and longer durations of morning stiffness in the nearest neighbour subset compared to the distribution in the entire representation cohort.

##### Ranking of features

Plots in Figure 10 and in the appendix S1 Appendix. Similarity provide insights into the nearest neighbour attribution mechanism on an individual patient level. Using the method described in subsubsection 2.3.1, we ranked the features by global importance in the cohort. We found that overall both DAS scores and the number of swollen joints are the most important for the similarity assessment for continuous features (Table 5). Similarly, high duration of morning stiffness and gender are the top-2 categorical features for the similarity assessment (Table 4).

**Table 4.**
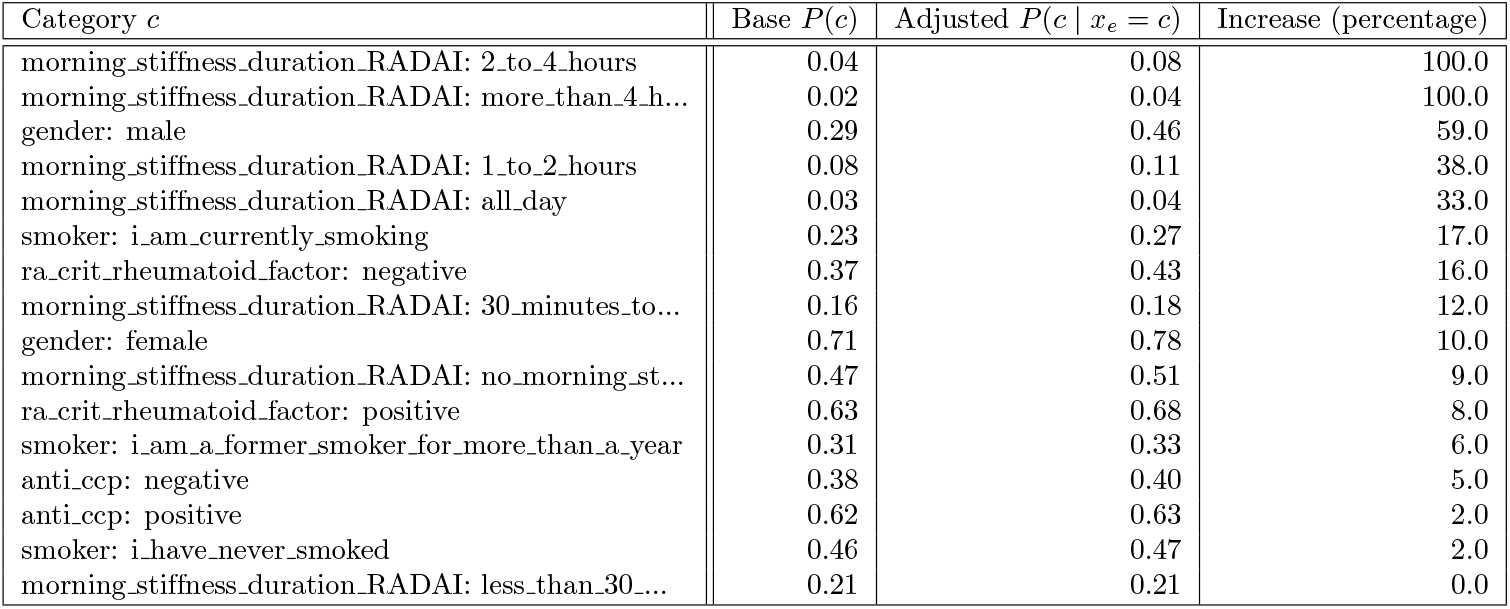
Similarity metric: contribution of categorical features. Empirical probability of a category *c* versus adjusted probability, given that the data point is in the subset of nearest neighbours *𝒩*_*e*_ of a datapoint *x*_*e*_ with the same category *c*. The increase in the adjusted probability reflects the importance of a given category in the similarity assessment. Longer durations of morning stiffness and gender have the strongest impact on the similarity assessment.

**Table 5.** Similarity metric: contribution of continuous features. Average absolute distance (AAD) and standardised AAD between the feature value of a test embedding *e*_*p,t*_ and the mean feature value within its nearest neighbours *𝒩*_*e*_. The features are ordered by standardised AAD. We see that the two DAS and the number of painful joints are taken into account the most during the similarity assessment.

| Feature | AAD | Standardised AAD |
| --- | --- | --- |
| asdas_score | 0.25 | 0.24 |
| das283bsr_score | 0.35 | 0.25 |
| n_painfull_joints_28 | 2.05 | 0.41 |
| n_painfull_joints | 2.40 | 0.43 |
| crp | 5.35 | 0.46 |
| n_swollen_joints | 2.14 | 0.50 |
| bsr | 8.03 | 0.50 |
| mda_score | 0.73 | 0.56 |
| n_enthesides | 1.42 | 0.58 |
| joints_type | 8.05 | 0.61 |
| haq_score | 0.46 | 0.65 |
| pain_level_today_RADAI | 1.91 | 0.71 |
| activity_of_rheumatic_disease_today_RADAI | 1.89 | 0.71 |
| hb | 0.97 | 0.72 |
| height_cm | 6.73 | 0.73 |
| weight_kg | 12.05 | 0.76 |

##### Clinical relevance of findings

Our analysis of patient similarity suggested that the impact of smoking on disease parameters varies among patients. Genetic association studies showed that smoking is only associated with an increased risk of developing RA in people carrying the shared epitope genes in the HLA-DR locus, but not in current smokers without these RA risk genes [22]. While it is known that smoking negatively affects treatment response and disease severity in both RA and axSpA [23–26], it would be interesting to know if this is the same in all patients or if genetic background plays a similarly important role in the impact of smoking on disease.

## Conclusion

In this work, we propose DAS-Net, a multitask neural network-based model for transforming heterogeneous rheumatic disease registry data into comparable patient representations and predicting future disease activity. When predicting future DAS, DAS-Net outperformed all non-temporal baseline models that discarded or oversimplified most of the patient history.

Our model design included attention layers that aided in explaining the importance of the different visits and parts of the patient’s history in outcome prediction. It showed that our model uses recent information but still attributes significant weight to older events and that the model attributes the majority of the weight to the clinical measures. This pattern gets stronger as the amount of available history increases and the model performance improves for longer medical histories.

Moreover, the predictive power of the nearest neighbour approach on the model’s latent representations showed that our model is well suited to transform heterogeneous electronic health records into comparable representations. One possible extension for our model would be to explicitly incorporate a clustering loss in the training objective [27] to further improve the patient similarity framework.

Lastly, the results of the three different analyses of feature importance (feature attribution via SHAP, attention weights and case-based similarity) are in concordance with clinical expert knowledge ([28], [29], [30]). Past disease activity scores were consistently the strongest predictors in all three analyses and gender and rheumatoid factor stood out as important features for the similarity assessment. Consistent with these findings, low disease activity, including low CRP/BSR levels, and low HAQ levels have also been associated with good future outcomes in patients with RA in previous studies [31, 32]. Similarly, autoantibody status and gender have been described before as predictors of outcomes in RA patients [32–34].

Overall, our study demonstrates promising results towards developing an explainable clinical decision support system for retrieving similar patients and predicting their disease progression while considering the different disease management strategies that worked best for similar patients. Such a CDSS would be especially useful for managing complex chronic diseases. It could help find optimal management strategies faster by assessing which strategy worked best for similar patients.

## Data Availability

Data are owned by a third party, the Swiss Clinical Quality Management in Rheumatic Diseases (SCQM) foundation and may be obtained after approval and permission from SCQM.

https://www.scqm.ch/

## Data and code availability

The code developed for the analysis is available on the following GitHub repository

https://github.com/uzh-dqbm-cmi/scqm.

## Acknowledgments

The authors thank the patients and caregivers who made the study possible, as well as the clinicians who collected the data. A list of rheumatology offices and hospitals that are contributing to the SCQM registries can be found on www.scqm.ch/institutions. The SCQM is financially supported by pharmaceutical industries and donors. A list of financial supporters can be found on www.scqm.ch/en/partners/. The authors thank Almut Scherer for her feedback on the manuscript.

## Supporting information

### 3.3 S1 Table. Dataframes and features

**Table S 1.** Name of input features in the different types of patient records in the SCQM database.

| Dataframe | demographics | clinical measures | medications | PROM |
| --- | --- | --- | --- | --- |
| Features | date_of_birth,<br>gender,<br>date_first_symptoms,<br>date_diagnosis | date,<br>weight_kg,<br>das283bsr_score,<br>asdas_score,<br>n_swollen_joints,<br>n_painfull_joints,<br>bsr,<br>n_painfull_joints_28,<br>height_cm,<br>crp,<br>hb,<br>n_enthesides,<br>mda_score,<br>joints_type,<br>anti_ccp,<br>ra_crit_rheumatoid_factor,<br>smoker,<br>haq_score | medication_generic_drug,<br>medication_drug_classification,<br>medication_dose,<br>date,<br>is_start | date,<br>pain_level_today_RADAI,<br>morning_stiffness_duration_RADAI,<br>activity_of_rheumatic_disease_today_RADAI |

### 3.4 S2 Table. Description of continuous clinical measure features

**Table S 2.** Mean, standard deviation and missingness of the continuous clinical measure features.

|  | mean | std | missing (%) |
| --- | --- | --- | --- |
| weight_kg | 72.78 | 16.03 | 30.21 |
| das283bsr_score | 3.02 | 1.40 | 14.45 |
| asdas_score | 2.34 | 1.03 | 84.63 |
| n_swollen_joints | 2.54 | 4.18 | 4.94 |
| n_painfull_joints | 3.42 | 5.51 | 18.70 |
| bsr | 16.33 | 15.91 | 0.98 |
| n_painfull_joints_28 | 2.99 | 4.86 | 14.36 |
| height_cm | 167.06 | 9.23 | 34.72 |
| crp | 6.56 | 11.33 | 26.74 |
| hb | 13.80 | 1.36 | 36.64 |
| n_enthesides | 1.31 | 2.40 | 78.62 |
| mda_score | 1.68 | 1.33 | 0.31 |
| haq_score | 0.77 | 0.71 | 40.16 |

### 3.5 S3 Table. Description of categorical clinical measure features

### 3.6 S4 Table. Description of continuous medication features

**Table S 4.** Mean, standard deviation and missingness of continuous medication features

|  | mean | std | missing (%) |
| --- | --- | --- | --- |
| medication_dose | 253.32 | 459.76 | 0.25 |

**Table S 3.** Distribution of categorical clinical measure features.

|  | smoker | anti_ccp | ra_crit_rheumatoid_factor | joints_type |
| --- | --- | --- | --- | --- |
| i_have_never_smoked (%) | 16.18 | NaN | NaN | NaN |
| i_am_a_former_smoker_for_more_than_a_year (%) | 11.05 | NaN | NaN | NaN |
| i_am_currently_smoking (%) | 7.71 | NaN | NaN | NaN |
| positive (%) | NaN | 36.34 | 51.44 | NaN |
| negative (%) | NaN | 22.40 | 30.07 | NaN |
| 28.0 (%) | NaN | NaN | NaN | 65.36 |
| 44.0 (%) | NaN | NaN | NaN | 22.39 |
| 68.0 (%) | NaN | NaN | NaN | 12.20 |
| missing (%) | 65.06 | 41.26 | 18.49 | 0.05 |

### 3.7 S5 Table. Description of categorical medication features

**Table S 5.** Distribution of categorical medication features.

|  | medication_generic_drug | medication_drug_classification | is_start |
| --- | --- | --- | --- |
| methotrexate (%) | 23.34 | NaN | NaN |
| prednisone (%) | 17.32 | NaN | NaN |
| rituximab (%) | 10.26 | NaN | NaN |
| adalimumab (%) | 8.09 | NaN | NaN |
| etanercept (%) | 6.68 | NaN | NaN |
| infliximab (%) | 5.77 | NaN | NaN |
| leflunomide (%) | 5.70 | NaN | NaN |
| sulfasalazine (%) | 5.09 | NaN | NaN |
| Other (%) | 3.53 | NaN | NaN |
| golimumab (%) | 3.40 | NaN | NaN |
| hydroxychloroquine (%) | 2.95 | NaN | NaN |
| tocilizumab (%) | 2.69 | NaN | NaN |
| abatacept (%) | 2.15 | NaN | NaN |
| certolizumab (%) | 2.15 | NaN | NaN |
| secukinumab (%) | 0.89 | NaN | NaN |
| bDMARD (%) | NaN | 42.68 | NaN |
| csDMARD (%) | NaN | 37.40 | NaN |
| steroid (%) | NaN | 17.35 | NaN |
| tsDMARD (%) | NaN | 2.23 | NaN |
| yes (%) | NaN | NaN | 54.96 |
| no (%) | NaN | NaN | 45.04 |
| missing (%) | 0.00 | 0.35 | 0.00 |

### 3.8 S6 Table. Description of continuous PROM features

**Table S 6.** Mean, std and missingness of continuous PROM features.

|  | mean | std | missingness |
| --- | --- | --- | --- |
| pain_level_today_RADAI | 3.01 | 2.59 | 0.57 |
| activity_of_rheumatic_disease_today_RADAI | 3.14 | 2.57 | 0.75 |

### 3.9 S7 Table. Description of categorical PROM features

**Table S 7.** Distribution of categorical PROM features.

|  | morning_stiffness_duration_RADAI |
| --- | --- |
| no_morning_stiffness (%) | 45.86 |
| less_than_30_minutes (%) | 23.87 |
| 30_minutes_to_1_hour (%) | 14.36 |
| 1_to_2_hours (%) | 7.10 |
| 2_to_4_hours (%) | 3.78 |
| all_day (%) | 2.31 |
| more_than_4_hours (%) | 1.83 |
| missing (%) | 0.89 |

### 3.10 S8 Table. Description of continuous demographic features

**Table S 8.**
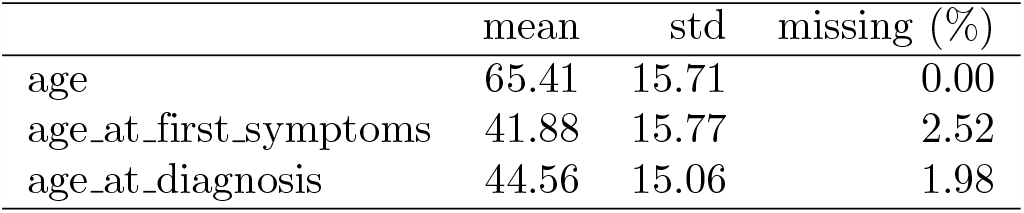
Mean, standard deviation and missingness of demographic features.

|  | mean | std | missing (%) |
| --- | --- | --- | --- |
| age | 65.41 | 15.71 | 0.00 |
| age_at_first_symptoms | 41.88 | 15.77 | 2.52 |
| age_at_diagnosis | 44.56 | 15.06 | 1.98 |

### 3.11 S9 Table. Description of categorical demographic features

**Table S 9.**
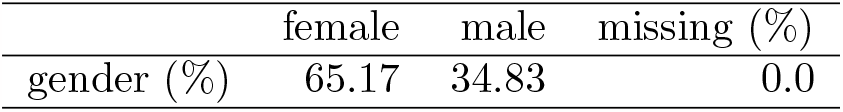
Distribution of categorical demographic features.

|  | female | male | missing (%) |
| --- | --- | --- | --- |
| gender (%) | 65.17 | 34.83 | 0.0 |

### 3.12 S1 Appendix. Similarity

The plots in Figure S 1 and Figure S 2 show additional t-SNE visualisations of patient representations. In each of the figures, the larger dot represents a randomly selected patient, and the triangle their nearest neighbours as computed by our algorithm.

**Figure S1.**
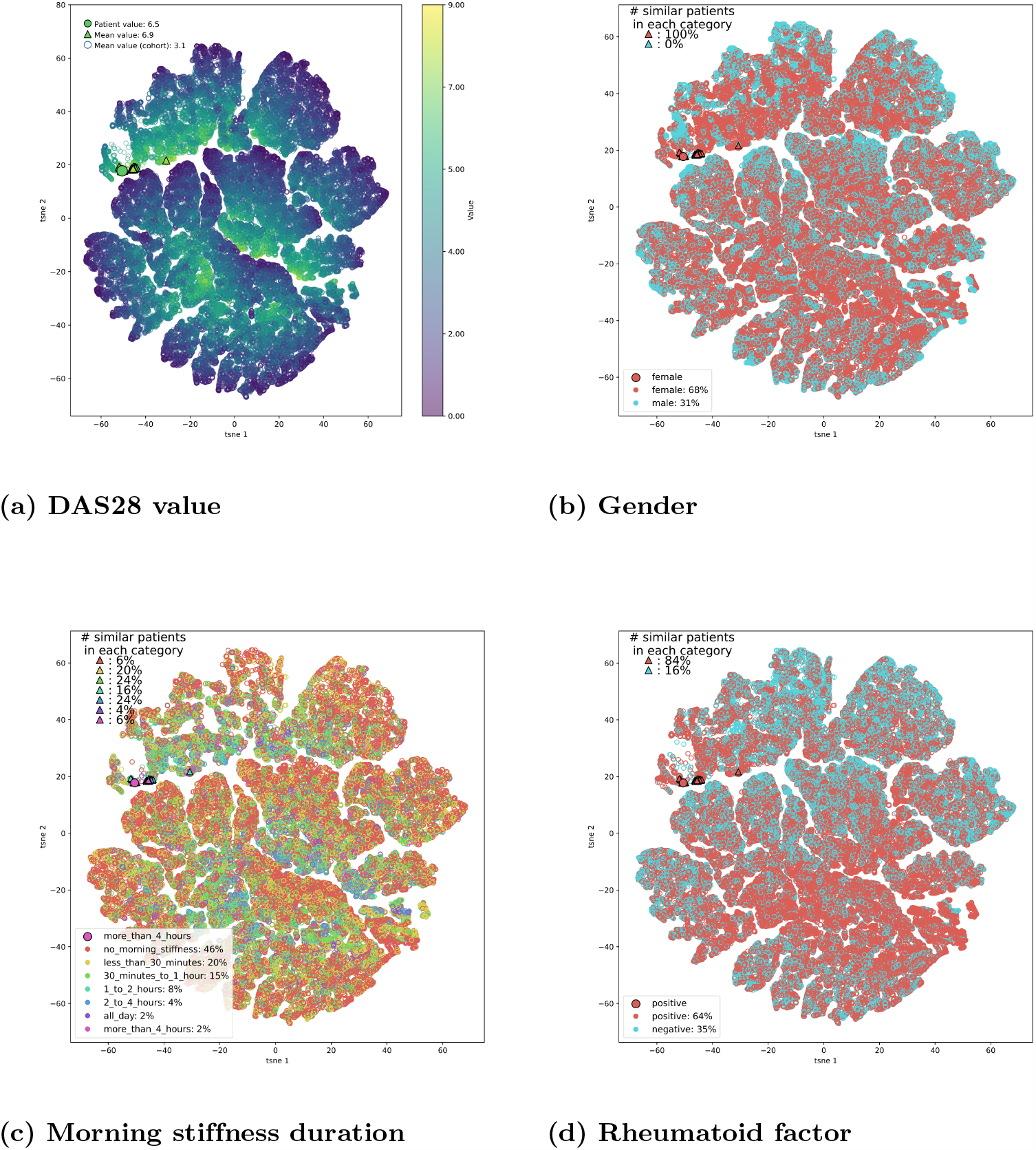
The DAS28 score and gender show a high level of consistency among the closest neighbors of this patient. Duration of morning stiffness and rheumatoid factors also show slight distribution shifts within the subsets of nearest neighbours.

**Figure S2.**
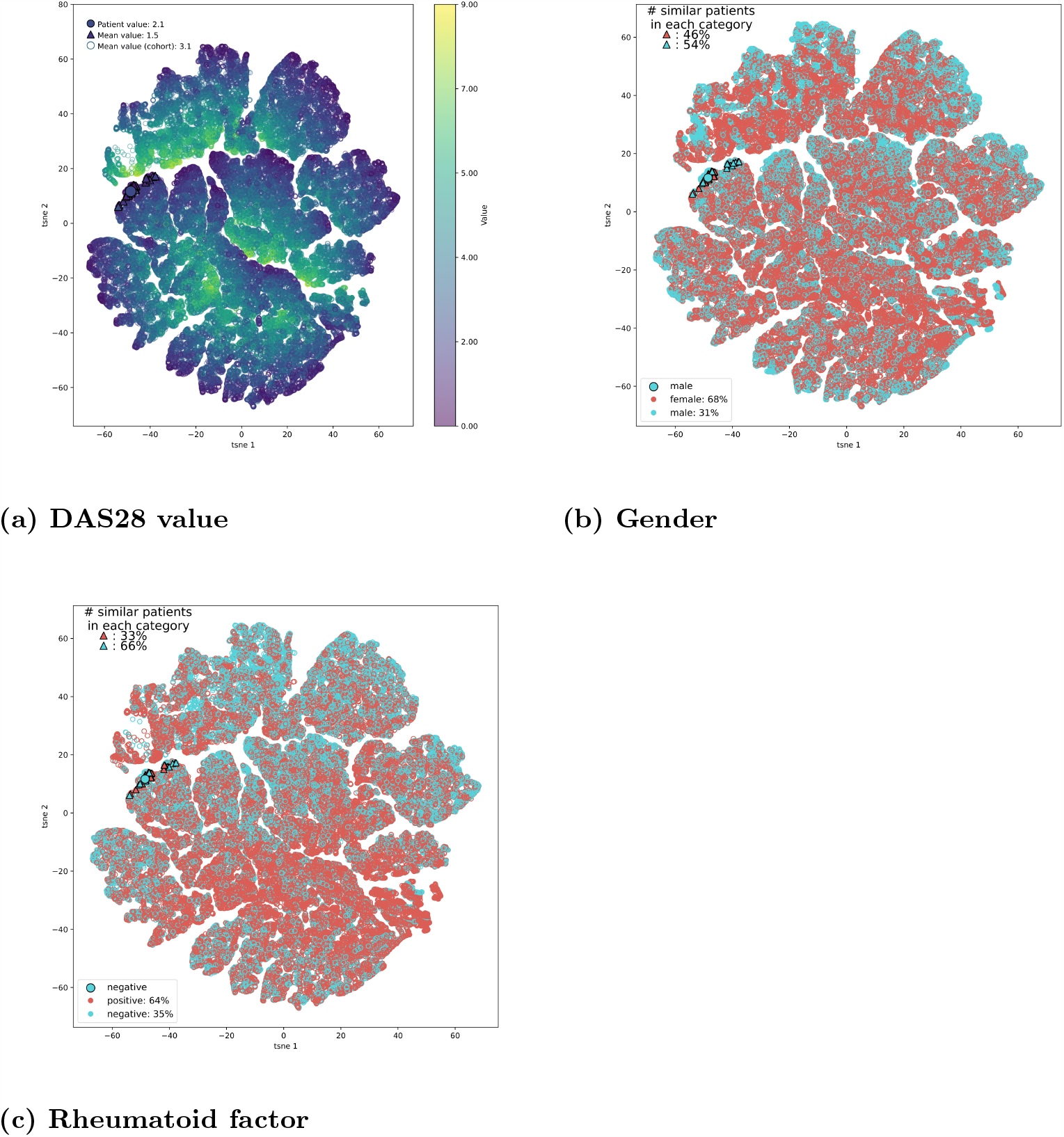
The patient’s rheumatoid factor status and their low DAS28 value can be observed among nearest neighbours subset as well.

## References

1. Chronic inflammation and your joints - Harvard Health;. Available from: https://www.health.harvard.edu/pain/chronic-inflammation-and-your-joints.

2. Kingsmore KM, Puglisi CE, Grammer AC, Lipsky PE. An introduction to machine learning and analysis of its use in rheumatic diseases. Nature Reviews Rheumatology. 2021;17(12):710–730.

3. Uitz E, Fransen J, Langenegger T, Stucki G. Clinical quality management in rheumatoid arthritis: putting theory into practice. Rheumatology. 2000;39(5):542–549.

4. The DAS28 score — NRAS — Disease Activity Score;. Available from: https://nras.org.uk/resource/the-das28-score/.

5. ASDAS calculator - ASAS;. Available from: https://www.asas-group.org/instruments/asdas-calculator/.

6. Hochreiter S, Schmidhuber J. Long short-term memory. Neural computation. 1997;9(8):1735–1780.

7. Bahdanau D, Cho K, Bengio Y. Neural machine translation by jointly learning to align and translate. arXiv preprint arXiv:14090473. 2014;.

8. Allam A, Dittberner M, Sintsova A, Brodbeck D, Krauthammer M. Patient similarity analysis with longitudinal health data. arXiv preprint arXiv:200506630. 2020;.

9. Karim MR, Beyan O, Zappa A, Costa IG, Rebholz-Schuhmann D, Cochez M, et al. Deep learning-based clustering approaches for bioinformatics. Briefings in bioinformatics. 2021;22(1):393–415.

10. Lundberg SM, Lee SI. A unified approach to interpreting model predictions. Advances in neural information processing systems. 2017;30.

11. Choi E, Bahadori MT, Schuetz A, Stewart WF, Sun J. Doctor ai: Predicting clinical events via recurrent neural networks. In: Machine learning for healthcare conference. PMLR; 2016. p. 301–318.

12. Montani S, Striani M. Artificial intelligence in clinical decision support: a focused literature survey. Yearbook of medical informatics. 2019;28(01):120–127.

13. Norgeot B, Glicksberg BS, Trupin L, Lituiev D, Gianfrancesco M, Oskotsky B, et al. Assessment of a deep learning model based on electronic health record data to forecast clinical outcomes in patients with rheumatoid arthritis. JAMA network open. 2019;2(3):e190606–e190606.

14. Lee S, Kang S, Eun Y, Won HH, Kim H, Lee J, et al. Machine learning-based prediction model for responses of bDMARDs in patients with rheumatoid arthritis and ankylosing spondylitis. Arthritis Research & Therapy. 2021;23:1–12.

15. Kalweit M, Walker UA, Finckh A, Müller R, Kalweit G, Scherer A, et al. Personalized prediction of disease activity in patients with rheumatoid arthritis using an adaptive deep neural network. PLoS One. 2021;16(6):e0252289.

16. Aletaha D, Smolen JS. Outcome Measurement in Rheumatoid Arthritis: Disease Activity. Rheumatoid Arthritis. 2009; p. 225–230.

17. Vaswani A, Shazeer N, Parmar N, Uszkoreit J, Jones L, Gomez AN, et al. Attention is all you need. Advances in neural information processing systems. 2017;30.

18. Loshchilov I, Hutter F. Decoupled weight decay regularization. arXiv preprint arXiv:171105101. 2017;.

19. Shapley L. Value for n-person games, contributions to the theory of games (Kuhn, HW, Tucker, AW Eds.). 307–317. Ann Math Stud. 1953;28:275–293.

20. Orange DE, Blachere NE, DiCarlo EF, Mirza S, Pannellini T, Jiang CS, et al. Rheumatoid arthritis morning stiffness is associated with synovial fibrin and neutrophils. Arthritis & Rheumatology. 2020;72(4):557–564.

21. Van der Maaten L, Hinton G. Visualizing data using t-SNE. Journal of machine learning research. 2008;9(11).

22. Padyukov L, Silva C, Stolt P, Alfredsson L, Klareskog L. A gene–environment interaction between smoking and shared epitope genes in HLA–DR provides a high risk of seropositive rheumatoid arthritis. Arthritis & Rheumatism: Official Journal of the American College of Rheumatology. 2004;50(10):3085–3092.

23. Masdottir B, Jonsson T, Manfreosdóttir V, Víkingsson A, Brekkan Á, Valdimarsson H. Smoking, rheumatoid factor isotypes and severity of rheumatoid arthritis. Rheumatology. 2000;39(11):1202–1205.

24. Hyrich K, Watson K, Silman A, Symmons DP, Register BB. Predictors of response to anti-TNF-α therapy among patients with rheumatoid arthritis: results from the British Society for Rheumatology Biologics Register. Rheumatology. 2006;45(12):1558–1565.

25. Ciurea A, Scherer A, Weber U, Exer P, Bernhard J, Tamborrini G, et al. Impaired response to treatment with tumour necrosis factor α inhibitors in smokers with axial spondyloarthritis. Annals of the rheumatic diseases. 2016;75(3):532–539.

26. Villaverde-García V, Cobo-IbÁñez T, Candelas-Rodríguez G, Seoane-Mato D, del Campo-Fontecha PD, Guerra M, et al. The effect of smoking on clinical and structural damage in patients with axial spondyloarthritis: a systematic literature review. In: Seminars in arthritis and rheumatism. vol. 46. Elsevier; 2017. p. 569–583.

27. Xie J, Girshick R, Farhadi A. Unsupervised deep embedding for clustering analysis. In: International conference on machine learning. PMLR; 2016. p. 478–487.

28. Albrecht K, Zink A. Poor prognostic factors guiding treatment decisions in rheumatoid arthritis patients: a review of data from randomized clinical trials and cohort studies. Arthritis research & therapy. 2017;19(1):1–8.

29. Favalli EG, Biggioggero M, Crotti C, Becciolini A, Raimondo MG, Meroni PL. Sex and management of rheumatoid arthritis. Clinical reviews in allergy & immunology. 2019;56:333–345.

30. Ishikawa Y, Terao C. The impact of cigarette smoking on risk of rheumatoid arthritis: a narrative review. Cells. 2020;9(2):475.

31. Burkard T, Williams RD, Vallejo-Yagüe E, Hügle T, Finckh A, Kyburz D, et al. Prediction of sustained biologic and targeted synthetic DMARD-free remission in rheumatoid arthritis patients. Rheumatology Advances in Practice. 2021;5(3):rkab087.

32. Schlager L, Loiskandl M, Aletaha D, Radner H. Predictors of successful discontinuation of biologic and targeted synthetic DMARDs in patients with rheumatoid arthritis in remission or low disease activity: a systematic literature review. Rheumatology. 2020;59(2):324–334.

33. Hamann PD, Pauling JD, McHugh N, Shaddick G, Hyrich K. Predictors, demographics and frequency of sustained remission and low disease activity in anti-tumour necrosis factor–treated rheumatoid arthritis patients. Rheumatology. 2019;58(12):2162–2169.

34. Vallejo-Yagüe E, Pfund JN, Burkard T, Clair C, Micheroli R, Möller B, et al. Lower odds of remission among women with rheumatoid arthritis: A cohort study in the Swiss Clinical Quality Management cohort. Plos one. 2022;17(10):e0275026.

